# Statistical methods for batch screening of input populations by stage and group in COVID-19 nucleic acid testing

**DOI:** 10.1101/2020.04.02.20050914

**Authors:** Cheng Yuan Yuan

## Abstract

**Purpose:** To screen for COVID-19 patients in immigration using minimal nucleic acid testing (NAT).

**Methods:** In the first phase, nasopharyngeal swab samples from the inbound population were numbered and grouped. The samples in the group were mixed together, and a NAT test was performed. When the test result is negative, it means that everyone in the group is not infected and the screening of the group is complete. When the test results were positive, the group moved on to the second stage. In the second stage, all samples in the positive group will be tested individually for NAT.

**Results:** The advantages and considerations of the method are discussed. Prevalence in the incoming population was a determinant of the sample size within the group. The lower the incidence, the larger the sample size within the group, the higher the savings in NAT and testing costs.

**Conclusion:** This method has significant efficiency and cost advantages in COVID-19 screening. It can also be used to screen other populations, such as community populations and people at high risk of infection, etc.

## Background

Since the WHO declared COVID-19 (Corona Virus Disease 2019) a global pandemic on March 11, 2020, ^[1]^ the number of Chinese arrivals has reached 600,000 by March 15, 2020, ^[2]^ and 44 new confirmed cases have been detected from the arriving population. ^[3-7]^ Starting from March 19, 2020, cities such as Shenzhen, Guangzhou, Shanghai, and Beijing have implemented the policy of nucleic acid testing (NAT) for all inbound persons. ^[8-11]^

## Method

The formula used is as follows:

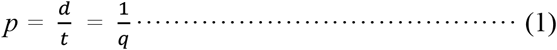

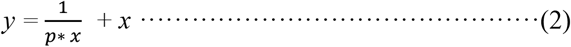

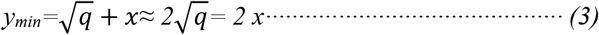

R software (Version 3.6.3) code to calculate y is as follows:

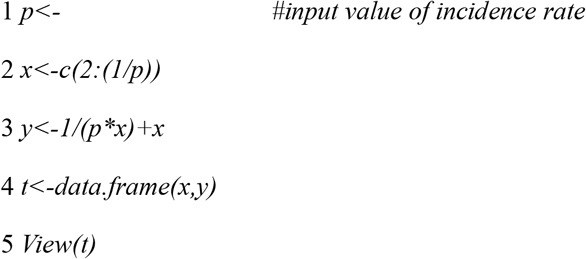

where *p* is the incidence of the incoming population, *d* is the number of confirmed cases in the incoming population, *t* is the number of concurrent incoming population, *q* is the number of incoming persons bringing one COVID-19 patient, *y* is the number of NATs, *x* is the number of samples per group, and *y*_*min*_ is the minimum number of NATs. According to equation (1), when *d* =44, *t* =60,000, *p* =0.7333 (1/10,000), *q* =13,637. The incidence in the inbound population was 0.7333 (1/10,000). On average, for every 13,637 entries, one COVID-19 patient was brought in. In order to identify the patient, a COVID-19 fluorescence *RT-PCR* test had to be performed on 13,637 entry persons. At 160 *RMB* per test, the cost of 13,637 NATs is 2.18 million *RMB*. In order to achieve the same detection effect with the lowest number of NATs, a staged, batch screening approach is recommended (Figure 1).

**Figure 1.**
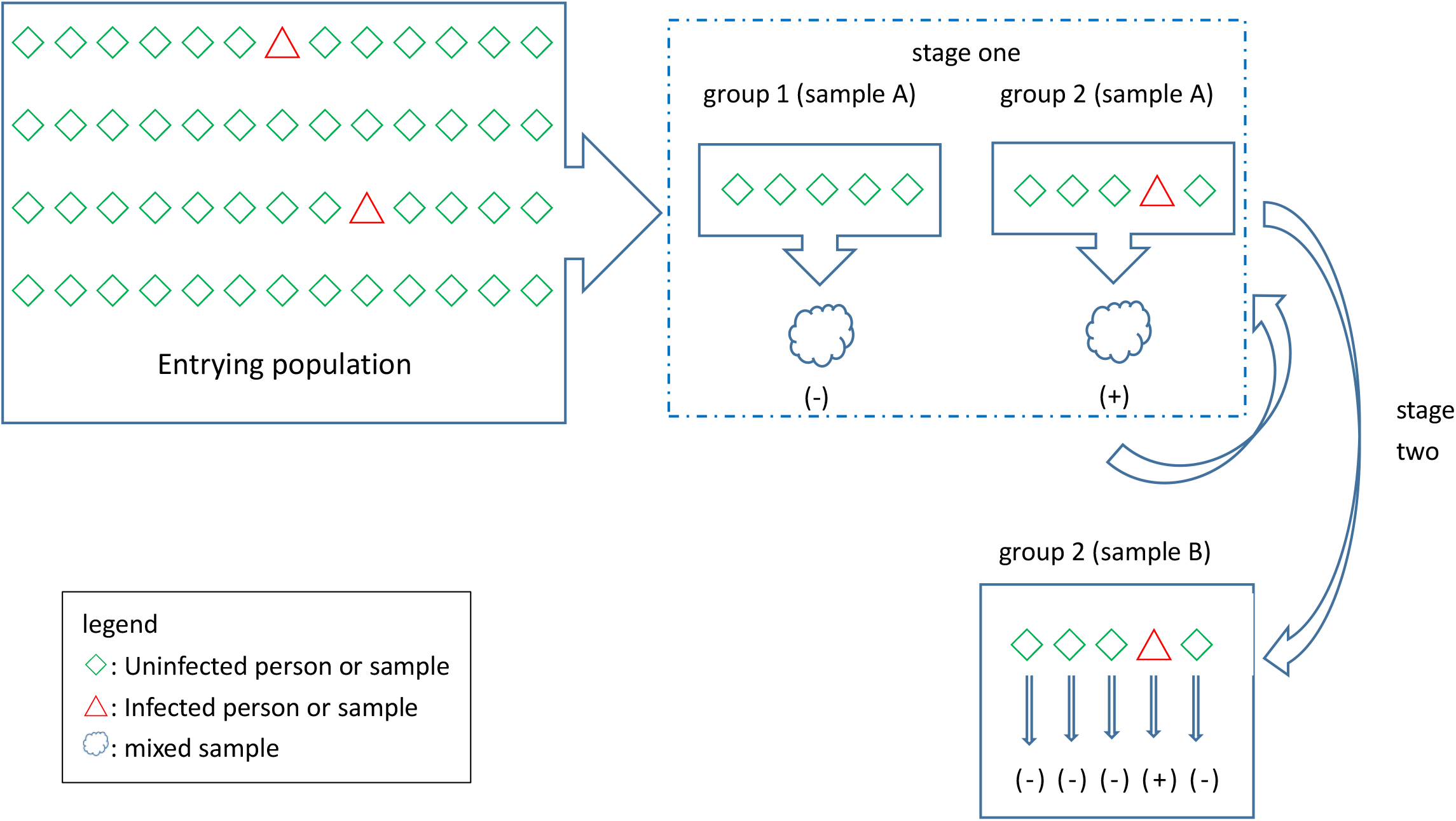
Schematic diagram of the process of phased batch screening in groups.

In the first stage, all arrivals are numbered, grouped, and each *x* is grouped. In the same group, a double nasopharyngeal swab was taken from each person, called sample A and sample B, and all sample A in the same group was mixed together for a single NAT test. When this test is negative, it means that everyone in the group is not infected and the test for that group is over. When a test result is positive, it means that at least one person in the group has a positive sample, and this group is called the “positive group”, and all samples in the positive group go on to the second stage of testing.

In the second stage, NAT was performed on all samples B in the positive group one by one to find out which sample caused the positive result and to screen the corresponding patients.

After two stages of bulk screening in groups, only *y* NAT is required, *q-y* NAT is saved.

We derive the minimum value of *y* by calculating x. The smaller the *p*, the larger the *q-y*, the more NATs can be saved and the greater the detection cost savings.

For example, assuming *p* was 0.733/100,000, then *q* was 13,637, according to formulas (1), (2), (3) and code of *R* software 3.63, when *x* was 117, the minimum value of *y* was 234; that was, 13,637 people were divided into 117 groups, each group of 117 people, the minimum value of NAT was 234, instead of 13,637. Each group of 117 people were taken double samples, with 234 nasopharyngeal swabs, as sample A and sample B, respectively. Each group had 117 samples A and 117 samples B, and all samples A mixed together for one NAT. When the test result was negative, it means that the test result of all 117 people was negative, the test of that group was over. When the test result was positive, it means that at least one of the 117 people tested positive. All samples B in the positive group underwent a second stage of testing. In the second stage, all samples B from the positive group were tested individually with NAT for a total of 117 tests. As a result, one patient was screened from 13,637, requiring only 234 NATs, saving 13,403 tests compared to 13,637 NATs. On average, each person entering the country pays only 2.8 *RMB* instead of 160 *RMB* for NAT, which equates to a savings of 2.14 million *RMB* in testing costs, a 98.3 per cent savings. The savings rate was higher if two or more patients were found in the same group, or if the method was repeated in the positive group. The cost of NAT was at least 960,000 *RMB*, based on the average of more than 6,000 test visits per day in Beijing. Using this method, Beijing can save 940,000 *RMB* per day in testing costs and 28.2 million *RMB* in 30 days. Based on the daily average of 120,000 passengers entering China through land ports, seaports and airports, the cost of testing can be saved by 18.83 million *RMB* in 30 days, which was *RMB* 565 million.

## Conclusion

The method is not limited to screening of inbound populations, but can also be used to screen community populations and at-risk populations. ^[12-27]^

It should be noted that p is the determinant of the number of people in each group *x*. The lower the *p*, the greater the value of *x*, the greater the value of *q-y* and vice versa (Table 1, Figure 2, Figure 3).

**Table 1:** Relationship between incidence, sample size, and savings rate

| $p(1/10,000)$ | $q$ | $x$ | $y_{min}$ | Percentage saved (%) |
| --- | --- | --- | --- | --- |
| 10.00 | 1,000 | 32 | 64 | 93.6 |
| 5.00 | 2,000 | 45 | 90 | 95.5 |
| 3.33 | 3,000 | 55 | 110 | 96.3 |
| 2.50 | 4,000 | 64 | 128 | 96.8 |
| 2.00 | 5,000 | 71 | 142 | 97.2 |
| 1.67 | 6,000 | 78 | 156 | 97.4 |
| 1.43 | 7,000 | 84 | 168 | 97.6 |
| 1.25 | 8,000 | 90 | 180 | 97.8 |
| 1.11 | 9,000 | 95 | 190 | 97.9 |
| 1.00 | 10,000 | 100 | 200 | 98.0 |
| 0.91 | 11,000 | 105 | 210 | 98.1 |
| 0.83 | 12,000 | 110 | 220 | 98.2 |
| 0.77 | 13,000 | 115 | 230 | 98.2 |
| 0.71 | 14,000 | 119 | 238 | 98.3 |
| 0.67 | 15,000 | 123 | 246 | 98.4 |

**Figure 2.**
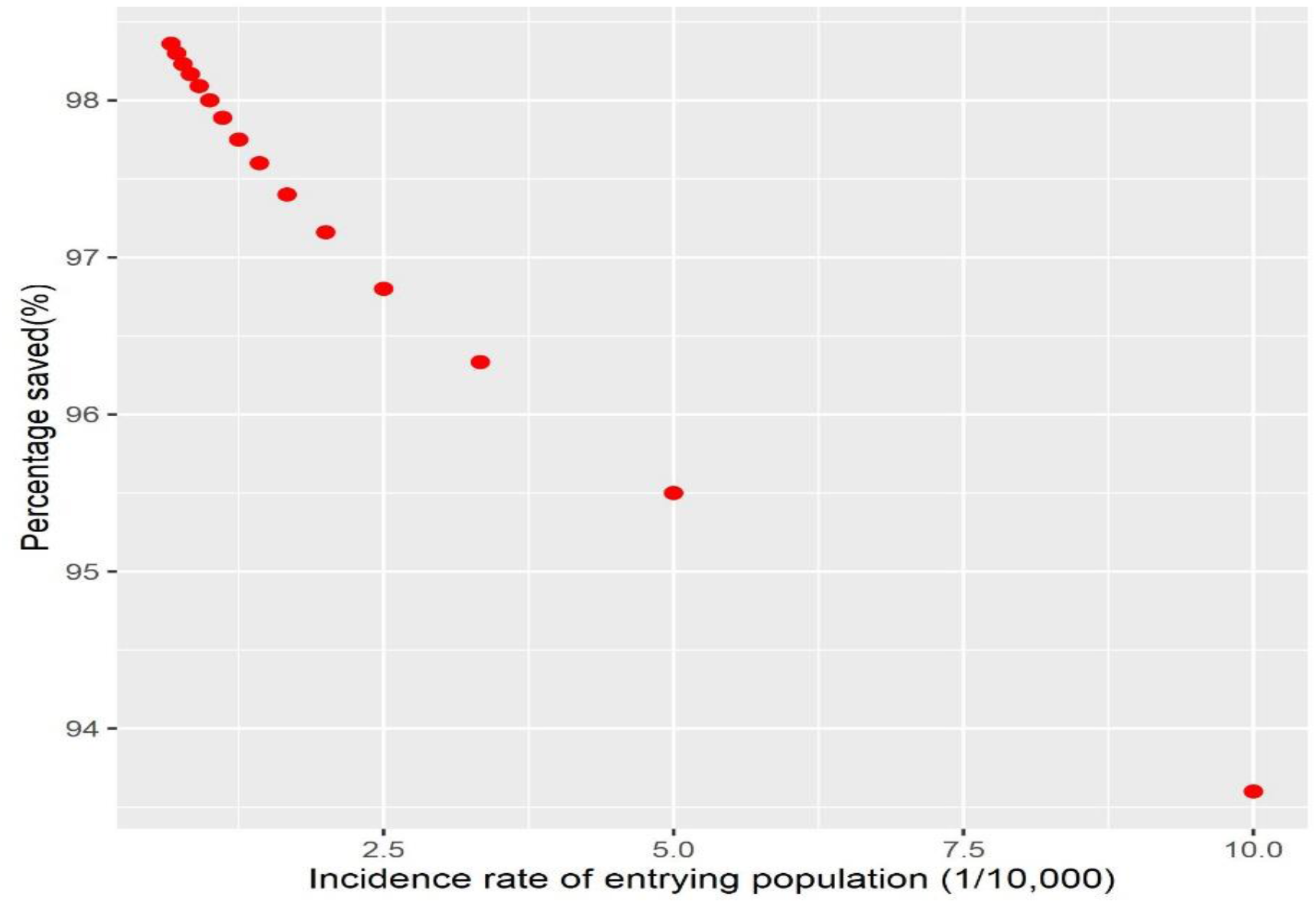
Scatterplot of incidence and percentage saved for the entrying population. The relationship between the incidence rate and the percentage of savings can be seen, with the lower the incidence rate, the greater the percentage of savings.

**Figure 3.**
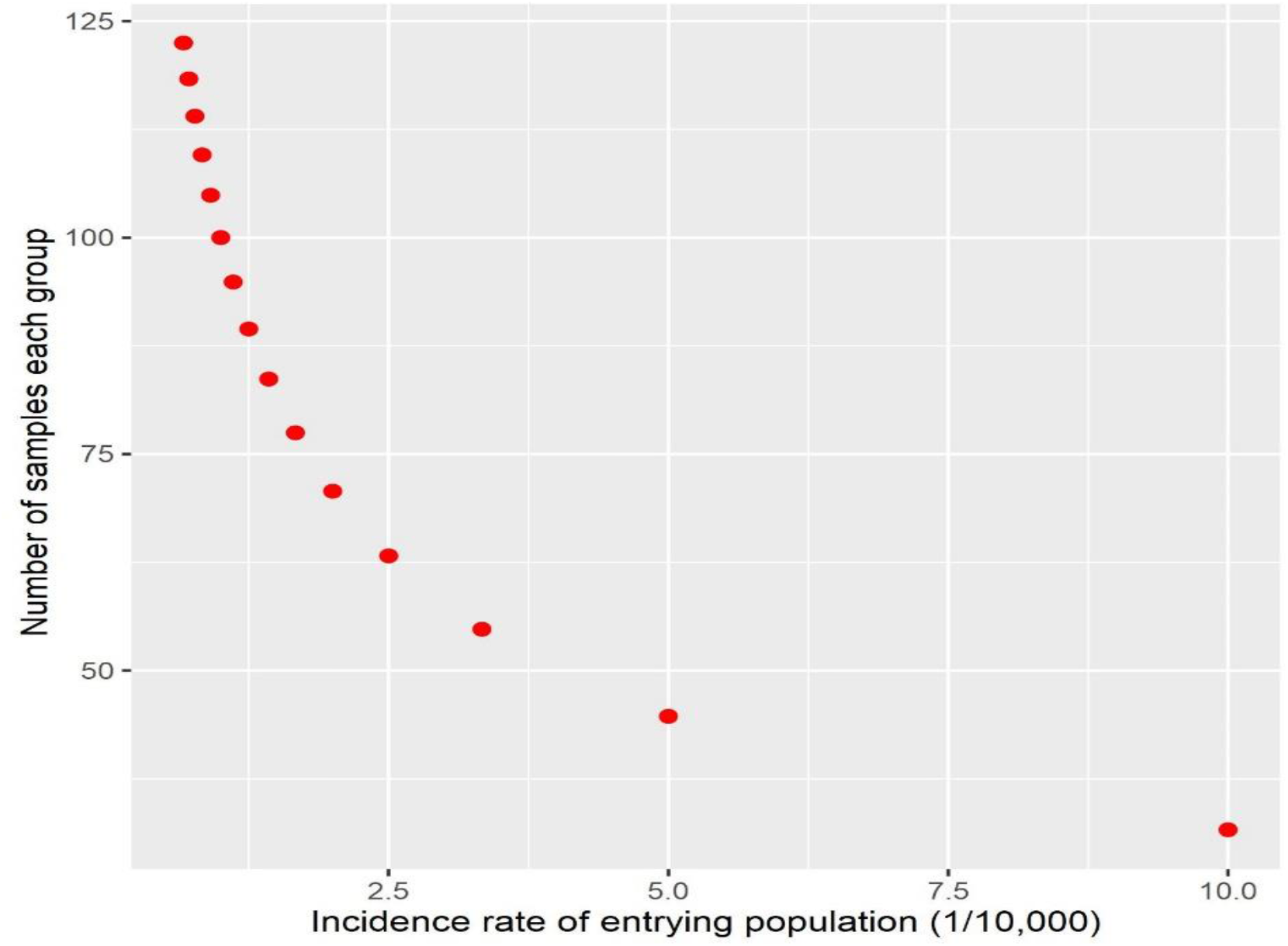
Scatterplot of incidence rates in the incoming population versus sample size per group. It can be seen that the incidence in the inbound population is related to the sample size per group, the lower the incidence, the larger the sample size per group, the more NATs can be saved.

There are many factors that affect *p*, such as international situation, country, city, observation period, customs policy, and the characteristics of the population entering the country. Sometimes these factors cause large fluctuations in *p* and require timely monitoring of *p* and adjustment of *x* according to Equation (2). when *p* fluctuates within a certain range, *x* should be adjusted according to the maximum value of *p*.

## Data Availability

All data, models, and code generated or used during the study appear in the submitted article.

## Competing Interest Statement

The authors declare no competing interests.

## Funding Statement

There is no external funding, and there is no conflict of interest.

## Author Declaration

All relevant ethical guidelines have been followed; any necessary IRB and/or ethics committee approvals have been obtained, and details of the IRB/oversight body are included in the manuscript.

Yes

All necessary patient/participant consent was obtained, and the appropriate institutional forms were archived.

Yes

I understand that all clinical trials and any other prospective interventional studies must be registered with an ICMJE-approved registry, such as ClinicalTrials.gov. I confirm that any such study reported in the manuscript has been registered and the trial registration ID is provided (note: if posting a prospective study registered retrospectively. Please provide a statement in the trial ID field explaining why the study was not registered in advance).

Yes

I have followed all appropriate research reporting guidelines and uploaded the relevant EQUATOR Network research reporting checklist(s) and other pertinent materials as supplementary files, if applicable.

Yes

## Notes

### Competing Interest Statement

The authors have declared no competing interest.

### Clinical Trial

NCT12345678

